# Cost-utility analysis of primary HPV testing through home-based self-sampling in comparison to visual inspection using acetic acid for cervical cancer screening in East district, Sikkim, India, 2023

**DOI:** 10.1101/2024.03.03.24303673

**Authors:** Roopa Hariprasad, Bhavani Shankara Bagepally, Sajith Kumar, Sangeeta Pradhan, Deepsikka Gurung, Harki Tamang, Arpana Sharma, Tarun Bhatnagar

## Abstract

**Introduction:** Primary Human Papilloma Virus (HPV) testing offers higher sensitivity and specificity over Visual Inspection using Acetic acid (VIA) in cervical cancer screening. Self-sampling is a promising strategy to boost participation and reduce disparities. However, concerns about the initial costs hinder HPV testing adoption in low and middle-income countries. This study assesses the cost-utility of home-based HPV self-sampling versus VIA for cervical cancer screening in India

**Methods:** A cross-sectional study was conducted in East district, Sikkim, India, comparing the costs and utility outcomes of population-based cervical cancer screening through VIA and primary HPV screening through self-sampling. Cost-related data were collected from April 2021 to March 2022 using the bottom-up micro-costing method, while utility measures were collected prospectively using the EuroQoL-5D-5L questionnaire. The utility values were converted into quality-adjusted life days (QALDs) for an 8-day period. The willingness to pay threshold (WTP) was based on per capita GDP for 2022. If the calculated Incremental Cost-Effectiveness Ratio (ICER) value is lower than the WTP threshold, it signifies that the intervention is cost-effective.

**Results:** The study included 95 women in each group of cervical cancer screening with VIA & HPV self-sampling. For eight days, the QALD was found to be 7.977 for the VIA group and 8.0 for the HPV group. The unit cost per woman screened by VIA and HPV self-testing was ₹1,597 (US$ 19.2) and ₹1,271(US$ 15.3), respectively. The ICER was ₹-14,459 (US$ −173.6), which was much below the WTP threshold for eight QALDs, i.e. ₹ 4,193 (US$ 50.4).

**Conclusion:** The findings support HPV self-sampling as a cost-effective alternative to VIA. This informs policymakers and healthcare providers for better resource allocation in cervical cancer screening in Sikkim.

## Introduction

Cervical cancer ranks as the second most prevalent cancer among women in India. Globocan estimates for 2020 showed 123,907 incident cases, and 77,348 cases succumbed to this disease [1]. In 2016, the Government of India, through the Ministry of Health and Family Welfare, implemented an operational framework with the goal of facilitating the screening and early detection of three preventable cancers: oral, breast, and cervical cancer [2]. Although visual inspection using the acetic acid (VIA) screening method adopted in the program for cervical cancer is evidence-based, there are concerns about the feasibility of its implementation in the field and beneficiary satisfaction [3,4]. Further, Human Papilloma Virus (HPV) testing exhibits higher sensitivity (98%) and specificity (90.6%) than VIA [sensitivity 31.6% and specificity 87.5%] [5].

In accordance with World Health Organization’s (WHO) recommendation, high-income countries have transitioned from cytology-based screening to primary HPV testing [6]. Research has demonstrated that self-collected samples for HPV testing by women yield comparable results to clinician-collected cervical samples [7,8]. Nevertheless, the implementation of HPV programs in low- and middle-income countries is hindered by the upfront financial burden associated with the adoption of this approach despite its evident advantages over VIA. A systematic review on the cost-effectiveness of all cervical cancer screening methods in low- and middle-income countries revealed that HPV testing through self-sampling is the most cost-effective to screen cervical cancer [9]. In the context of India, a single modeling study has examined the cost-effectiveness of three cervical cancer screening strategies: VIA, Pap smear, and HPV DNA [10]. However, to obtain more precise and trustworthy insights, empirical data is required. By utilizing real-world data, we can gather more reasonable information on the actual costs, effectiveness, and outcomes of cervical cancer prevention strategies when implemented within the Indian healthcare system. Such empirical studies are crucial for enhancing the applicability and validity of findings, empowering policymakers to make well-informed decisions regarding resource allocation for cervical cancer prevention programs in India.

Sikkim, a northeastern state in India, has piloted primary HPV screening through self-sampling in the East district to explore the challenges in implementation and planning to scale up to all districts based on the results and availability of funds for the same. In order to consider HPV testing as the primary screening of cervical cancer in the State, a detailed economic analysis using contextual data is needed. An essential factor in evaluating the validity of a new screening program is its ability to demonstrate economic efficiency while also surpassing the physical, psychological, and societal harms stemming from the associated testing and screening procedures. Thus, the current study was planned to assess the cost-utility of home-based HPV self-sampling compared to VIA for cervical cancer screening among women over 30 years in the East district of Sikkim state.

## Methods

### Study design and setting

We did a cross-sectional study in the East district, Sikkim, India, in which cost-related data was collected retrospectively, while the data for utility measures was collected prospectively. Fig 1 illustrates the study workflow. It includes estimates on the cost of population-based cervical cancer screening through VIA (facility-based) under the National Program for Prevention and Control of Non-Communicable Diseases (NP-NCD) program and primary HPV screening through self-sampling (home-based) from a disaggregated societal perspective. This perspective considers costs estimated at both the health provider/system level and the individual level, specifically focusing on the direct expenses borne by women undergoing screening in the form of out-of-pocket payments.

**Fig 1:**
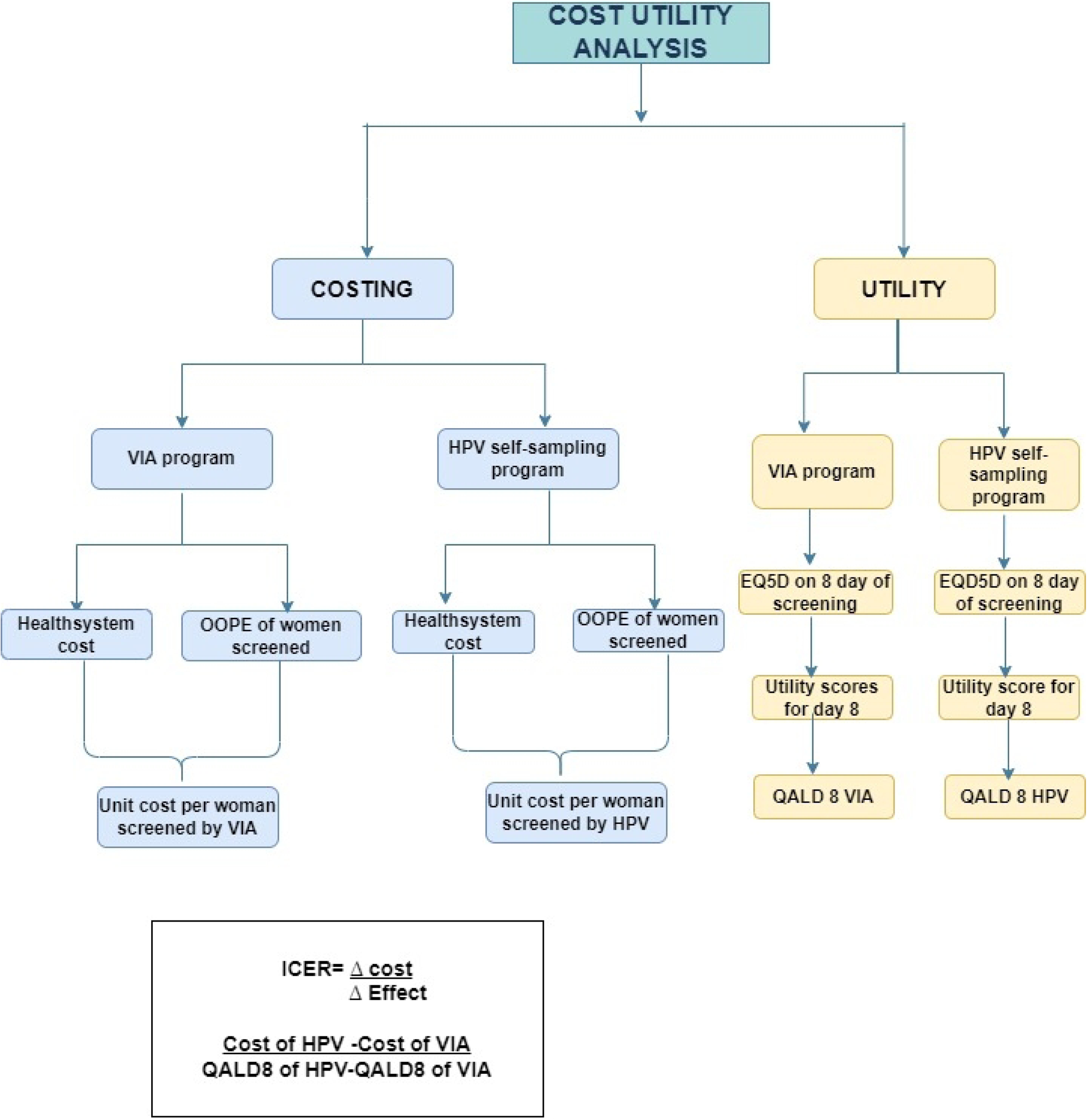
Illustration of the Study Work Flow.

Under the NP-NCD, VIA is performed on the eligible population (women aged 30 years and older) by the Community Health Officer (CHO) at Health and Wellness Centres (HWCs). If they are found to be VIA positive, women are referred to PHC for confirmation by Primary Health Centre (PHC) Medical Officer. In the primary HPV screening group, Accredited Social Health Activists (ASHAs) provide the sampling kits to women when they make home visits for either filling Community Based Assessment Checklist (CBAC) or Reproductive and Child Health (RCH) program-related activities. Community Health Workers in India are referred to as ASHAs. They are equipped with Information Education and Communication (IEC) materials, such as charts and videos, which demonstrate the correct procedure for collecting vaginal samples. The woman then collects the sample herself and hands over the sampler kit back to ASHA. The sample is transported to the HPV lab at the district hospital from PHC in an ambulance, which does a weekly visit to District Hospital for transportation of other contingencies like vaccines and medicines.

The detailed flow of the screening process in both groups is provided in Fig A of the supplementary file. The activities in both groups are the same once the woman is referred to PHC after screening. Hence, the unit cost is calculated only up to the screening process. Participants’ eligibility criteria for calculating the utility values were as follows; women aged 30 years or older who underwent screening using VIA at HWCs by CHOs, and women aged 30 years or above who underwent screening using HPV self-sampling at home were included in the study. Women who declined to provide consent were not included in the study. The reference period for costing was from April 2021 to March 2022.

Considering the mean EuroQol-5 dimension score (SD) of women after cervical cancer screening by VIA to be 0.89 (0.19) and the mean EQ-5D-5L score (SD) of women after HPV self-sampling to be 0.98 (0.19) [11] and power as 85%, alpha as 5% (two-sided), we calculated a sample size of 81 women in each group. Adjusting for a 15% attrition rate, the sample size was calculated to be 95 women in VIA and HPV groups. East district of Sikkim consists of eight PHCs (Two urban and six rural) and forty-three health and wellness centres (HWCs). HPV self-sampling (home-based) is implemented by the state govt in the East district in three PHC catchment areas (two urban and one rural). These HWCs selected for HPV screening do not perform VIA screening anymore. Out of the remaining thirty-two HWCs in the district, the best performing eleven HWCs in terms of number of women screened by VIA were selected.

### Data collection-costs

For cost analysis, costing related to human resources, space/building, furniture, equipment, consumables, IEC Material, stationery and overheads for both screening methods was obtained. Utility measures were calculated using EQ-5D-5L scores. Cost analysis in this study was undertaken using a bottom-up approach of micro-costing methods. All costs were reported for the reference year 2022 (financial year April 2021 to March 2022). All the resources used in screening for cervical cancer using VIA and HPV were identified, measured and valued. Data on capital and recurrent resources utilised to deliver cervical cancer screening services during the reference year were considered. Costing related to space/building, human resources, furniture, equipment, consumables, IEC Material, stationery and overheads for both screening methods were obtained from records and the data of the same were accesssed between 15^th^ January 2023 to 30^th^ April 2023.

For estimating the building cost, the current market rental price of a similar space through three key informant interviews was collected for each of the eleven HWCs and the HPV laboratory. The average of the three rental prices was considered. To allocate the cost specifically to the VIA screening activity, an apportioning factor was calculated considering the working time dedicated to VIA screening in a year relative to the total working hours of the healthcare facility.

To estimate the human resources cost, the investigator interviewed the study participants and healthcare persons using a structured questionnaire. The information related to salary/wages, perks and time spent on each of the activities of screening were collected. The human resources costs associated with screening were calculated using the apportioning factor derived from the time spent on activities.

The consumable cost was assessed using the cost, quantity and procurement date of consumables procured during the reference period for VIA and HPV tests by the state central procurement division. The details were obtained from their stock register and accounts division. For women screened by VIA, the Out of Pocket Expenditure (OOPE) accounted for direct costs associated with transportation to the facility, as well as any food expenses incurred during the screening period for both the women and their attendants, if accompanied. However, in the HPV group, it was not applicable as screening took place at home. The cost of overheads like water, maintenance, the electricity of HWCs and HPV testing lab was obtained at the facility level. The total annual cost incurred by each group was calculated by summing up the costs associated with various components. Unit cost was calculated by dividing the total cost of screening by the number of individuals screened in each group as unit cost per woman screened by VIA and unit cost per woman screened by HPV self-sampling

### Data collection-utility

EQ-5D-5L questionnaire was used for the personal interview [12]. Permission to use the paper, telephonic, and digital versions has been obtained from the EuroQol office. The questionnaire has five dimensions graded across five levels of problems. The tool comprises a descriptive questionnaire along with a visual analog scale (VAS) to assess perceived problems. These problem levels were assigned numerical codes ranging from 1 to 5, with each state represented by a 5-digit code. The questionnaire encompasses five dimensions, namely mobility, self-care, usual activities, pain/discomfort, and anxiety/depression, and investigates five problem levels, including the absence of problems, minor problems, moderate problems, significant problems, and extreme difficulties or inability to perform.

EQ-5D-5L was administered to all the Women undergoing cervical cancer screening by VIA at HWCs as well as women undergoing cervical cancer screening at home by HPV self-sampling between the period 15^th^ January 2023 to 30^th^ April 2023. Cervical cancer screening by VIA is performed at HWCs by trained CHOs as a part of the ongoing NP-NCD program. The questionnaire was administered after the screening procedure is complete and before the CHO informs the VIA result to the woman. For women undergoing HPV self-testing at home, it was administered after she collected the sample.

Permission from women to contact her telephonically after eight days was taken in the consent form. The EQ-5D-5L questionnaire was administered telephonically on the 8^th^ day of screening, and the status of symptoms was collected as per the satisfaction questionnaire.

### Data analysis

#### Scoring in EQ-5D-5L and calculation of QALD for 8 eight days

The EQ-5D-5L questionnaire responses from study participants were converted into a single index value using India-specific tariff values. This conversion process considered the health-related quality of life (HRQoL) across all five dimensions (mobility, self-care, usual activities, pain/discomfort, anxiety/depression) and assigned a weighted value to each health state. Once the EQ-5D-5L index value was obtained for each individual, it was multiplied by 8 (duration of interest) to calculate the QALDs. This calculation combines the individual’s health state utility value with the duration, reflecting the overall quality of life experienced during that period. The EQ-5D-5L VAS scale assesses the self-evaluated quality of life on a scale of 1 (worst imaginable health state) to 100 (best imaginable health state). Median quality of life with interquartile range was reported.

### Utility score

An EQ-5D-5L summary index was calculated by deducting the appropriate weights from 1, the value for full health (i.e., state 11111). For estimating the quality-of-life utility values, health states generated from the scoring on the EQ-5D-5L descriptive system was converted into utility scores using the India’s value set [13] (which is a collection of index values (weights) for all possible EQ-5D-5L states) was used for estimating the utility score.

### Incremental Cost-Effectiveness Ratio (ICER)

As per WHO guidance, the Willingness to Pay (WTP) threshold was determined based on the per capita GDP for the year 2022, amounting to ₹ 1,91,288 (US$ - 2297.7) per QALY. Then, for an eight (days period) QALDs, the WTP of ₹ 4193 (US$ 50.4) was considered. The Incremental Cost-Effectiveness Ratio (ICER) was then computed by dividing the difference in costs (incremental cost) between the two groups by the difference in the QALD8 scores. If the calculated ICER value is lower than the WTP threshold, it signifies that the intervention is cost-effective.

### Uncertainty analysis

A one-way sensitivity analysis was conducted to examine the effects of varying the number of individuals screened using both HPV & VIA methods. Additionally, the analysis included adjustments of all cost components with 10% and 25% variations, and the results were graphically presented.

Scenario analysis was performed to evaluate the potential impacts of different testing facility settings, including rural or urban locations. Furthermore, the analysis considered the reduction in HPV cost resulting from competitive pricing offered by manufacturers. We also explored the scenarios, where a healthcare provider at the facility conducted HPV testing, as well as the option of self-sampling at the facility. Statistical analysis was done using Epi-info version 7.2.1.0

All the costs are reported both in Indian National Rupees (INR) −₹ and United States Dollar (US$) (1 US$ = ₹83.2526). (https://www.exchangerates.org.uk/USD-INR-16_08_2023-exchange-rate-history.html)

### Ethical approval

Study proposal was approved by the Institute Human Ethics Committee of the ICMR-National Institute of Epidemiology, Chennai, India (Reference number: NIE/IHEC/A/202212-02). All the respondents were interviewed after obtaining written informed consent.

## Results

The baseline and screening attributes of women (95 in each group) interviewed in the VIA and HPV screening group are provided in Table 1. Both the HPV group and the VIA group were comparable in terms of age distributions, marital status, educational status, parity and age at first intercourse. However, there was a significant disparity in residence patterns, with VIA consisting of a higher proportion (99%) of participants residing in rural areas, while HPV predominantly comprised individuals living in urban regions (84%). There was a notable distinction in occupational profiles that emerged, with a greater proportion of homemakers (70%) observed in the VIA group, while the HPV testing group exhibited a more diverse range of occupations.

**Table 1:** Demographic characteristics of screened women in VIA and HPV group.

|  | Women screened by<br>VIA (n=95) | Women screened by<br>HPV (n= 95) |
| --- | --- | --- |
| **Age (years)** |  |  |
| Mean (SD) | 43.6 (±9.8) | 41.3 (±8.5) |
| Range (yrs) | 30-65 | 30-63 |
| **Residence n (%)** |  |  |
| Urban | 1 (1) | 80 (84) |
| Rural | 94 (99) | 15 (16) |
| **Education n (%)** |  |  |
| Literate | 89 (94) | 87 (92) |
| Illiterate | 6 (6) | 8 (8) |
| **Occupation n (%)** |  |  |
| Home maker | 66 (70) | 49 (51) |
| Others | 29 (30) | 46 (49) |
| **Marital status n (%)** |  |  |
| Ever married | 87 (91) | 88 (92) |
| Others | 8 (9) | 7 (8) |
| **Age at first intercourse** |  |  |
| <18 years | 17 (18) | 14 (15) |
| 18 years and above | 78 (82) | 81 (85) |
| **Parity** |  |  |
| [mean (SD)] | 2.44 ±1.5 | 2.05±1.2 |
| **Symptoms after screening** |  |  |
| (any) n (%) |  |  |
| Present | 70 (74) | 8 (9) |
| Absent | 25 (26) | 87 (91) |
| **Time taken to reach the facility** |  |  |
| ≤1 hour | 86 (90) | NA |
| More than one hour | 9 (10) |  |
| **Time spent on the screening test** |  |  |
| ≤ 30 min | 85 (89) | 95 (100) |
| > 30 min | 10 (11) | 0 |
| **Out of pocket expenditure (INR)** |  |  |
| Median {IQR } | 120 {60, 200} | NA |

Abbreviations- VIA: Visual Inspection using Acetic acid, HPV: Human Papilloma Virus, NA: Not
Applicable

After screening, it was observed that the VIA group reported a higher prevalence of symptoms (74%) compared to the HPV group (9%). There was a significant difference among the groups regarding time spent on screening. The out-of-pocket expenditure (OOPE) of women in the VIA group ranged from 0 to ₹ 6000 (US$ 0 to 72.1) with a median of ₹120 (IQR ₹ 60-200) (US$ 1.4, IQR), which. Time to reach the facility and OOPE did not apply to the HPV group since the screening was conducted in their homes.

The total annual expenditure on screening women for cervical cancer through VIA and HPV DNA tests amounted to ₹5,79,059 (US$ 6955.4) and ₹28,16,019 (US$ 33825), respectively. The details of the annual costs for both screening methods are provided in Table 2. The distribution of the total annual costs for VIA and HPV screening revealed that training (33%) and consumables (30%) comprised the largest proportion of overall expenses in the VIA group. Notably, in the HPV group, approximately 40% (₹11,29,706, US$ 13569.6) of the total cost was allocated to the purchase of consumables, followed by human resources, which accounted for 30% (₹ 8,55,848 US$ 10280.1) of the expenditure.

**Table 2:** Distribution of total annual cost by inputs for VIA and HPV based screening for cervical cancer during the financial year of 2021-22, Sikkim.

| Inputs | Annual cost in VIA group |  | Annual cost in HPV group |  |
| --- | --- | --- | --- | --- |
| | INR | US\$ | (INR) | US\$ |
| Space/Building | 8447 | 101.5 | 3,54,281 | 4255.5 |
| Furniture & Equipment | 12,111 | 145.5 | 1,60,859 | 1932.2 |
| Human resource | 1,52,803 | 1835.4 | 8,55,848 | 10280.1 |
| Consumables | 1,76,136 | 2115.7 | 11,29,706 | 13569.6 |
| IEC Material | - | - | 22,600 | 271.5 |
| Training and meetings | 1,91,000 | 2294.2 | 1,57,125 | 1887.3 |
| Overheads | 38,562 | 463.2 | 1,35,600 | 1628.8 |
| **Total cost** | **5,79,059** | **6955.4** | **28,16,019** | **33825** |
| No of tests | 392 |  | 2216 |  |
| **Unit cost per test** | **1,597** | **19.2** | **1,271** | **15.3** |

Abbreviations- VIA: Visual Inspection using Acetic acid, HPV: Human Papilloma Virus, IEC: Information
Education & Communication

During the study period, 392 women underwent VIA screening across eleven HWCs. These HWCs covered a population of 6,031 eligible women, resulting in a screening coverage of 6.5%. In the HPV group, 2,216 women were screened out of the 6,331 eligible women, yielding a screening coverage of 35%. The unit cost per woman screened by VIA and HPV was ₹ 1,597 (US$ 19.2) and ₹ 1,271 (US$ 15.3), respectively.

The comparison of EQ-5D-5L index scores of screened women in VIA and HPV groups is tabulated in Table 3. The mean EQ-5D-5L score was significantly higher in the HPV group (1.00±0) than in the VIA group (0.9903±0.02) (p<0.0001). On the eighth day of screening, the EQ-5D-5L scores remained significantly higher in the HPV group (1.000±0) than in the VIA group (0.9972±0.0118) (p=0.02). The overall health score on the day of screening was significantly higher in the HPV group (88.18±5.44) compared to the VIA group (84.4±6.41), (p<0.0001).

**Table 3:** EQ-5D-5L index scores of screened women in VIA and HPV group.

| EQ-5D-5L |  | VIA | HPV | p |
| --- | --- | --- | --- | --- |
| Utility score (0-1)<br>(mean score±SD) | Day 1 | 0.9903 ±0.0210 | 1.0000±0 | <0.0001 |
| | Day 8 | 0.9972±0.0118 | 1.0000 ±0 | 0.02 |
| Visual Analogue Score (1-100)<br>(mean score±SD) | Day 1 | 84.4 ±6.41 | 88.18±5.44 | <0.0001 |
| | Day 8 | 89.23 ±3.42 | 90.75 ±3.66 | 0.04 |

Abbreviations- VIA: Visual Inspection using Acetic acid, HPV: Human Papilloma Virus

None of the women in the HPV group complained of any deviation from normal in the five dimensions of the EQ-5D-5L. However, in the VIA group, 17% and 7% experienced pain and anxiety, respectively, on day one; on day eight, 3% reported anxiety. (Supplementary Table A)

For a duration of 8 days, the QALD was found to be 7.977 for the VIA group and 8 for the HPV group. Hence the ICER was ₹-14,459 (US$ 173.7). This is much below the WTP threshold for eight days, i.e., ₹ 4,193 (US$ 50.4).

The sensitivity analysis results for changes in the number of women screened using the VIA method is shown in Fig. 2A. As the number of screened women decreases, the ICER increases. When the number of screened women exceeds 549, HPV self-testing ceases to be more cost-effective than VIA. This implies that a screening coverage increase of 40% is required for VIA to be considered cost-effective than HPV self-testing.

**Fig 2A:**
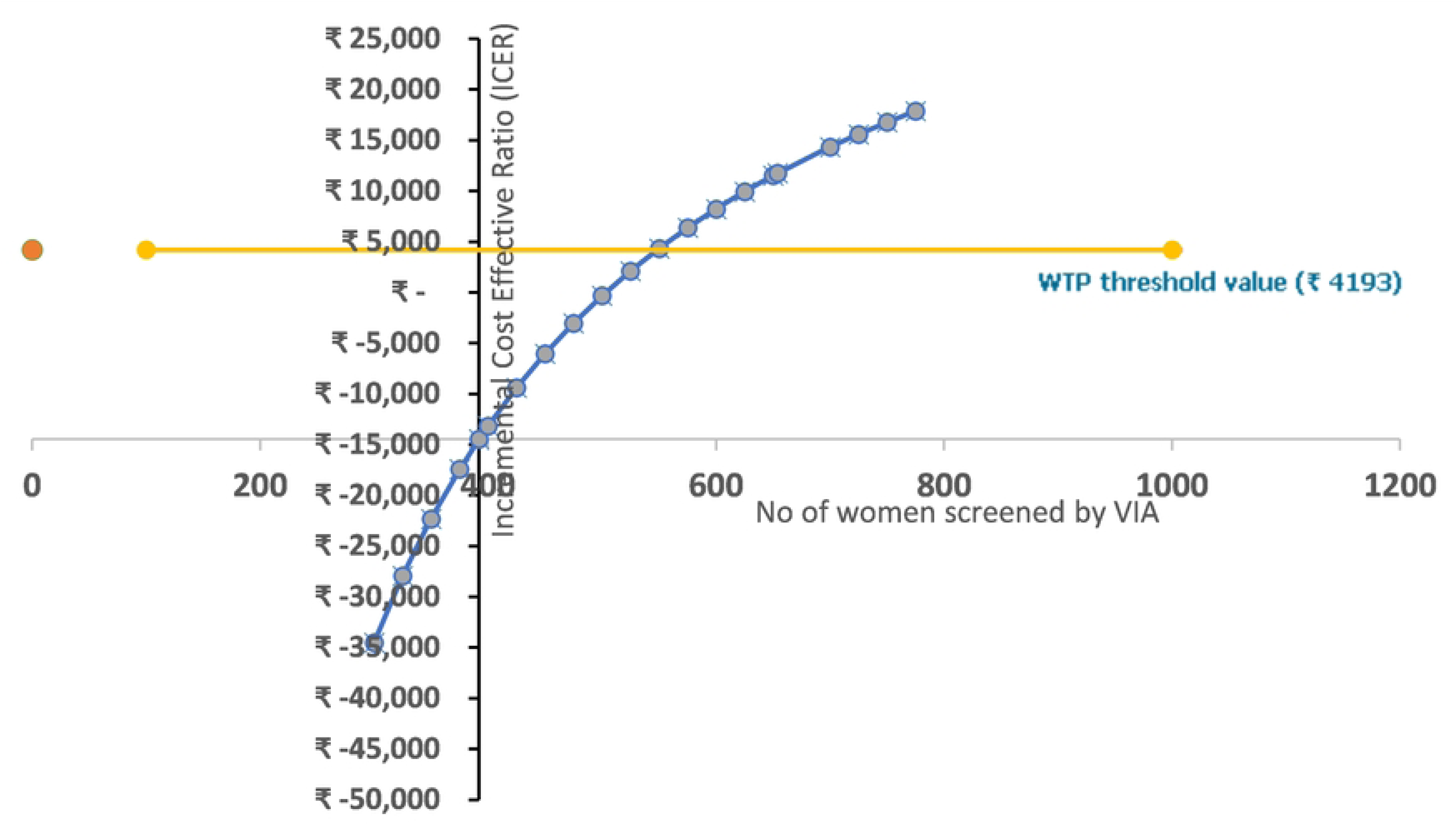
Sensitivity analysis: Change in ICER value with variation in the number of women screened by VIA.

In contrast, Fig. 2B demonstrates the impact of changes in the number of women screened by the HPV self-testing method on the ICER. As the number of women screened by HPV self-testing decreases, the ICER increases. Specifically, if the screened numbers fall below 1,665, HPV self-testing ceases to be cost-effective. However, even with a 25% lowering in screened numbers, the HPV self-testing method remains cost-effective.

**Fig 2B:**
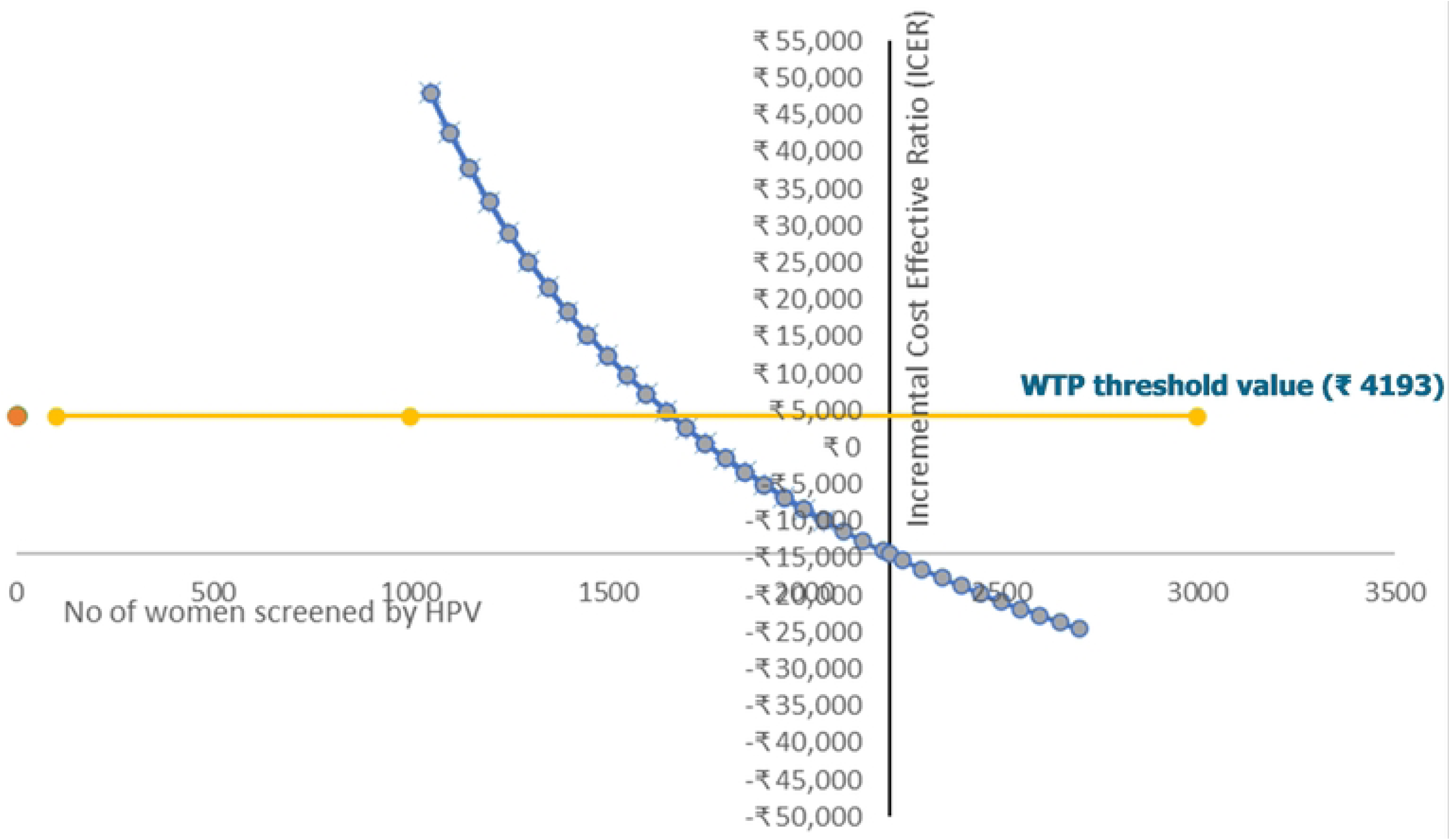
Sensitivity analysis: Change in ICER value with variation in the number of women screened by HPV.

The sensitivity analysis results for VIA screening (Fig. 3A) highlighted that training expenses and consumables had the highest impact on the ICER. Conversely, for HPV screening, consumables and human resources were found to have the greatest impact on the ICER (Fig 3B). ICER was below the WTP threshold for all the listed scenarios listed. (Supplementary Table B)

**Fig 3A:**
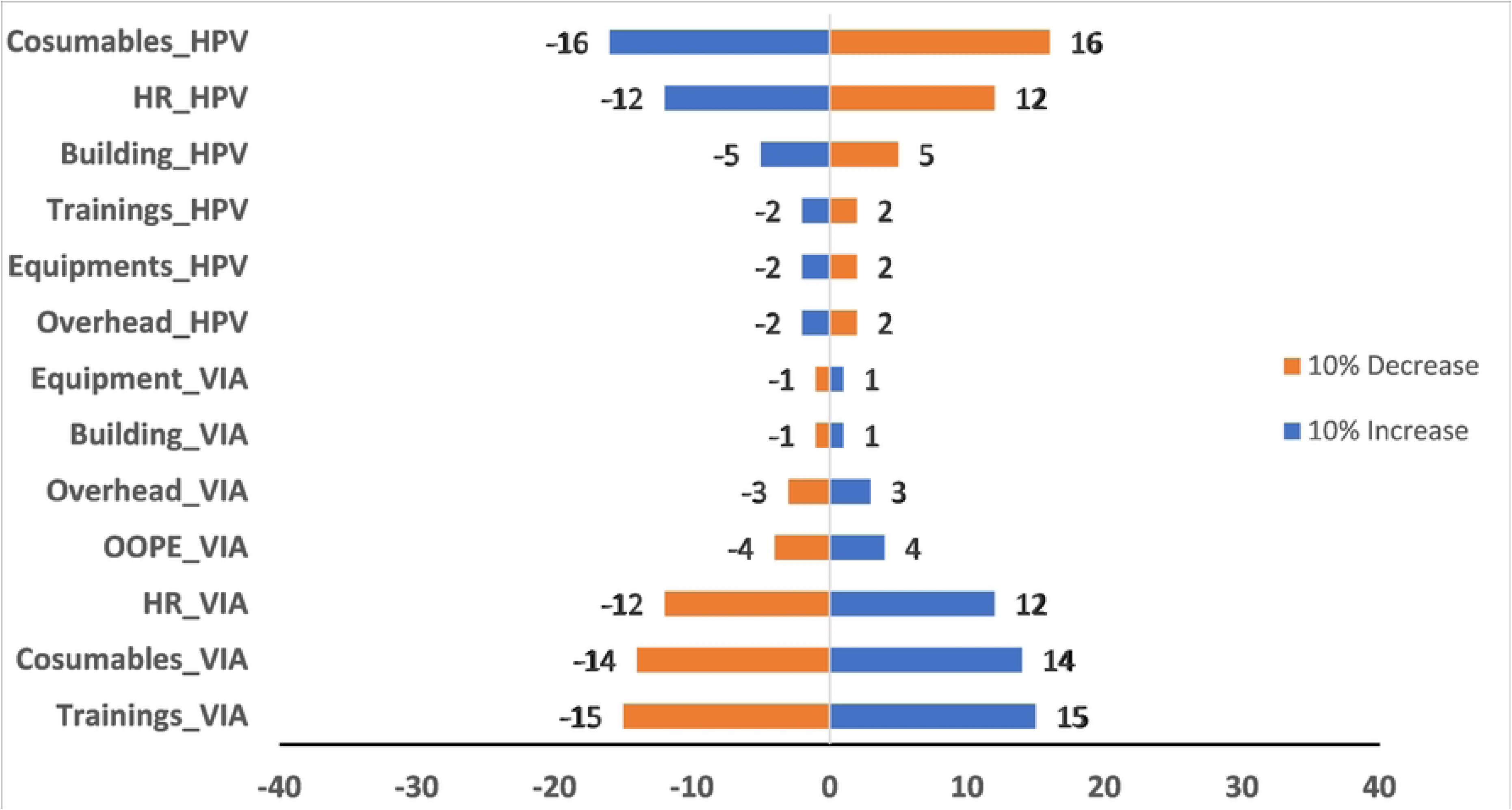
Sensitivity analysis; Key driving factors of ICER with 10% variation in item cost.

**Fig 3B:**
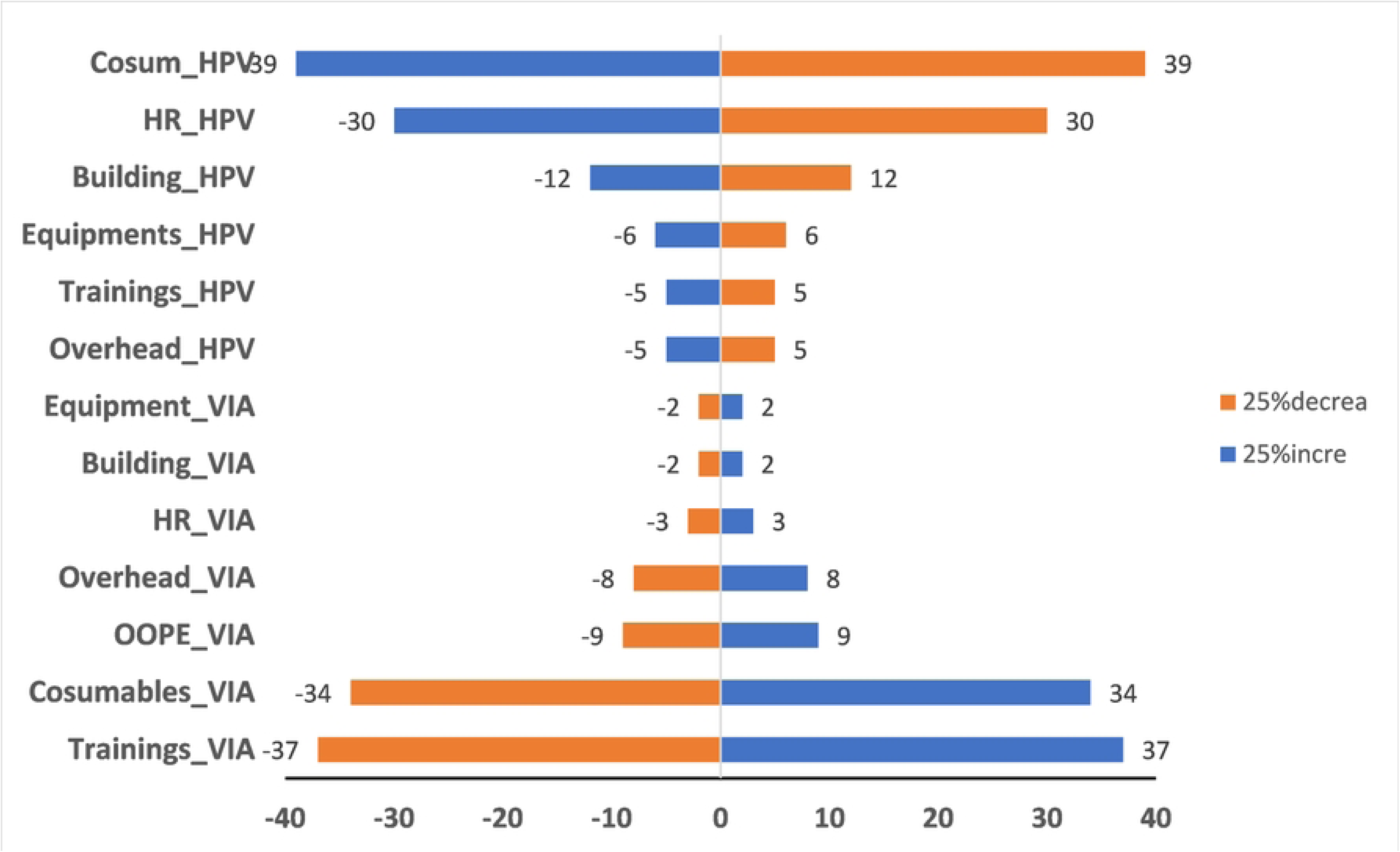
Sensitivity analysis; Key driving factors of ICER with 25% variation in item cost.

## Discussion

The findings of our study indicate that primary HPV screening through self-sampling is a cost-saving approach for cervical cancer screening, compared to the current recommendation of VIA for population-based screening in India. While the total cost of HPV screening may appear higher, the unit cost per test is substantially lower than VIA, primarily due to the higher screening coverage achieved with HPV screening. HPV self-sampling has been widely recognized for its ability to increase participation in cervical cancer screening as it overcomes various barriers; self-sampling effectively encourages more individuals to participate in regular screening programs, especially in low and middle-income countries [14,15]. Existing literature indicates that adopting self-sampling approaches can enhance screening participation among women and potentially lead to cost reductions [16]. According to a meta-analysis of 29 randomized controlled trials, the utilization of cervical cancer screening services was found to be twice as likely among women who underwent self-sampling compared to those who followed standard-of-care screening practices [17]. Women exhibit a preference for self-collection of samples within the convenience of their homes, as opposed to visiting healthcare facilities for screening [18]. It was found that there was a high level of compliance among women who opted for this self-collection method, with fewer operational challenges encountered during its implementation. Self-sampling presents a distinct opportunity to reach populations that are typically difficult to access, making it one of the most viable strategies for post-pandemic screening[19]. Self-sampling for HPV holds the promise of diminishing disparities in cervical screening participation within specific demographics and extending access to underserved and previously unscreened women [20,21].

Although VIA is known as a low-cost test and is recommended in low- and middle-income countries, the implementation of this subjective test is a major challenge. An Indian randomized controlled trial demonstrated that a single HPV test can effectively decrease the incidence of advanced cervical cancer and associated mortality. In contrast, VIA screening did not exhibit similar benefits in terms of cancer prevention and mortality reduction [22]. The significant hurdle to the widespread implementation of the more precise and efficient HPV screening method arises from the substantial upfront expenses required to establish screening programs. Although cost plays a vital role, it should not be the exclusive factor guiding decision-making. It is imperative to consider the well-being of women, along with their overall quality of life and satisfaction [23].

Our study findings also show significant differences in quality of life between women screened by HPV self-sampling who reported higher utility scores and a better quality of life compared to those who underwent VIA. The higher utility scores indicate that women screened through HPV self-sampling experienced better health-related quality of life, potentially due to reduced anxiety, pain and discomfort associated with the self-sampling approach. These results highlight the potential benefits of implementing HPV self-sampling as a preferred screening method, providing women with a more positive screening experience and ultimately contributing to improved health outcomes.

Previous studies have evaluated the cost-effectiveness of HPV self-sampling and VIA for cervical cancer screening [10, 24]. However, unlike these model-based studies, our study uniquely utilizes empirical data, marking the first instance of such an approach in India. An earlier model-based study in India that assessed the cost effectiveness of three screening strategies ie. Papanicolaou test, VIA and clinician based primary HPV testing for cervical cancer screening concluded VIA done every 5 years to be cost effective approach [10]. However, the study measured the health-related quality of life of cervical cancer patients rather than screened women to calculate the utility values. This approach failed to capture the immediate change in quality of life associated with the screening procedure itself. In addition, the study did not take into account the building cost of VIA screening as they reported data from camp-based approach.

Another study that evaluated cost effectiveness of same three screening strategies within a large cluster randomized trial from rural India (*Legood R et al., 2005*). The study dismissed HPV to be a cost-effective method for cervical cancer screening. It is important to note that this study accounted for all the costs in 2002, during which HPV testing was relatively new and prices were high, likely due to reduced competition in the market. Moreover, the study was conducted in a controlled settings of the trial that would vary from ‘real life’ situations. The ASPIRE randomized controlled trial included a cost-effectiveness study, which determined that community-based self-collected HPV testing is a cost-effective screening strategy [25].

Our study had few limitations. Building costs are typically higher in urban areas compared to rural areas. Differential implementation of HPV testing in urban areas and VIA testing in rural areas could have led to an overestimation of costs associated with the HPV testing strategy and an underestimation of costs associated with the VIA strategy. It is crucial to consider this limitation when interpreting the cost comparisons between the two screening methods. Another limitation of our study originates from the exclusive implementation of HPV screening within the East district of Sikkim [26]. The adoption of HPV screening specifically within urban area in the district has resulted in the inclusion of women who underwent VIA screening solely from rural PHC catchment regions. This specific sampling approach introduces a potential bias in the recruitment of participants, which may in turn limit the generalizability of the study findings.

In this study we have leveraged empirical data to evaluate the cost-utility of population-based self-collected HPV testing that is performed at women’s homes. The findings of this study provide a robust support for advocating the adoption and implementation of HPV self-sampling in India. The results indicate that the benefits obtained outweigh the associated costs, establishing it as a cost-effective approach within the provided threshold.. Further research following a similar framework in diverse settings across India is recommended. These studies would contribute to a more comprehensive understanding of the feasibility of HPV self-sampling as a primary screening method in various regional contexts.

## Data Availability

Deidentified (anonymized) data will be available on request

## Supplimentary file

a. Table A: EQ-5D-5L frequencies and proportions reported by dimensions and level
b. Table B: Comparison of ICER and unit cost for different cervical cancer screening scenarios
c. Fig A: Flow of screening process in HPV and VIA group

